# How knowledge shapes community stigma and social support for women seeking abortion in the Democratic Republic of Congo: A cross-sectional study

**DOI:** 10.64898/2026.06.18.26355933

**Authors:** Frédéric Etsou, Michée Kanda, Joseph Ngwanza, Bergitho Bokamba, Mike Mpoyi, Jean-Claude Mulunda, Nadia Lobo

**Affiliations:** Ipas DRC, Kinshasa, Democratic Republic of Congo; Institut Supérieur de Statistique, Kinshasa, Democratic Republic of Congo

**Keywords:** Social Support, Stigma, Reproductive Health, Abortion, Democratic Republic of Congo

## Abstract

**Background:** The Democratic Republic of Congo (DRC) bears one of the highest maternal mortality ratios globally (746 per 100,000 live births), with nearly 11% of deaths attributable to complications of unsafe abortion. Despite ratification of the Maputo Protocol and related national policies, access to safe abortion remains limited, largely due to entrenched stigma. Social support, encompassing emotional, informational, and instrumental assistance, is critical in shaping women’s abortion-seeking behaviors and health outcomes. This study examines the influence of community-level knowledge on stigma and social support for women seeking abortion care.

**Methods:** A cross-sectional survey was conducted from May 2024 to June 2024 among 1,715 adults in Kinshasa and North Kivu provinces. Analyses focused on a sub-sample of 574 respondents reporting familiarity with women who had undergone abortion. Structural Equation Modeling (SEM) was applied to estimate direct and indirect pathways linking community knowledge, stigma, and social support.

**Results:** Two core knowledge indicators, recognition of abortion as a safe medical procedure and awareness of legal conditions for access, were significantly associated with outcomes. A one-unit increase in knowledge corresponded to a 0.39-point increase in social support and a 0.19-point reduction in stigma. Enhanced knowledge promoted empathetic attitudes, reinforced practical support, and mitigated moralizing judgments toward women seeking abortion.

**Conclusions:** Strengthening community knowledge emerges as a strategic lever to reduce abortion-related stigma and enhance social support in the DRC. These findings underscore the importance of integrating stigma-reduction and knowledge-enhancement interventions into reproductive health programs to improve women’s access to safe and dignified abortion care.

**Plain-Language Summary:** In many communities, women who seek abortion face two big challenges: stigma (negative judgments and discrimination) and a lack of social support. These challenges often make it harder for women to get safe and respectful care.

This study looked at how knowledge about abortion, such as knowing that abortion can be safe medical procedure when performed by trained providers, and knowing the legal conditions under which it is allowed, affects stigma and social support in the Democratic Republic of Congo.

We used a survey, and a statistical method called structural equation modeling to understand the links between knowledge, stigma, and support. Our findings show that when community members have more accurate knowledge, women receive more support from their families and communities. For example, people are more willing to listen to them, help pay for care, or provide emotional encouragement. At the same time, increased knowledge reduces negative attitudes such as believing that women who have abortions are “bad mothers” or should be excluded from community life.

In short, better knowledge in communities leads to stronger support and less stigma. These results suggest that education and communication programs about abortion could play an important role in making communities more supportive and less judgmental toward women. Involving community leaders, health workers, and religious leaders in these efforts may help reduce stigma and improve access to safe abortion care in the DRC.

## 1. Background

The Democratic Republic of Congo (DRC) remains among the countries with the highest maternal mortality rates globally, estimated at 746 deaths per 100,000 live births according to the 2023–2024 Demographic and Health Survey (DHS). Abortion constitutes a critical public health issue in the DRC. Data from the 2024 Maternal, Perinatal, and Response Death Surveillance (SDMPR) report, produced by the National Reproductive Health Program, indicate that 11% of maternal deaths are attributable to abortion-related complications (1).

Access to safe abortion care represents a major public health challenge worldwide, particularly in the DRC. The World Health Organization (WHO) estimates that approximately 25 million unsafe abortions occur annually, accounting for nearly 45% of all abortions globally(2,3). Such unsafe procedures are associated with severe complications, including infections and hemorrhage, and contribute substantially to maternal mortality, particularly in resource-limited settings such as the DRC (4).

Despite the DRC’s ratification of the Maputo Protocol and the adoption of instruments such as the Standards and Guidelines for Comprehensive Woman-Centered Abortion Care (CAC), access to safe abortion remains constrained by unfavorable legal, sociopolitical, and cultural contexts. The operationalization of these frameworks is limited, and social support for women seeking abortion care remains inadequate. Consequently, many women continue to rely on clandestine or informal services (5–9). For example, in Kinshasa, it is estimated that over 146,000 abortions were performed in 2016, most outside formal health facilities (5).

Central to this issue is the concept of social support, defined as emotional, informational, and material assistance provided by family, professionals, or institutions, which plays a fundamental role in the care-seeking process for individuals pursuing abortion (9–12). Social support can influence the decision to undergo an abortion, the manner in which care is sought, and the overall experience of the procedure (10–13). Prior research identifies four primary dimensions of functional support: instrumental support, informational and appraisal support, emotional support, and companionship support (14).

However, in the DRC, patriarchal social norms, conservative religious beliefs, and cultural perceptions of sexuality and motherhood create an environment of high stigma surrounding abortion. This stigma limits the availability of support, silences women, and exposes them to unsafe care (6,15,16). Women experiencing pregnancies resulting from sexual violence, particularly in the eastern provinces, face compounded stigma related to both the assault and abortion, increasing their isolation (16–18).

According to the 2023–2024 community assessment conducted by Ipas DRC in the provinces of Kinshasa and North Kivu, social support for women seeking abortion is lower in North Kivu (mean score = 2.4/5) compared to Kinshasa (mean score = 3.1/5) on the Received Social Support Scale (R3S). Conversely, stigma is slightly lower in North Kivu (mean score = 2.84/5) than in Kinshasa (mean score = 2.87/5). Evidence suggests that social support significantly mitigates depression among women who have undergone abortion (19), highlighting its potential as a critical leverage point for facilitating access to stigmatized services such as safe abortion.

For communities to effectively support women seeking abortion care, they must possess a robust knowledge base regarding the legally permissible circumstances for abortion, understand the safeguards surrounding medical procedures, and be aware of the locations or providers authorized to offer such services. Access to accurate information thus constitutes a key determinant in building social support(20). However, knowledge alone is insufficient; in highly stigmatized contexts, women often pursue clandestine routes even when abortion is legally permitted (21).

This research addresses the needs of women and girls broadly, including those living in the eastern region of the Democratic Republic of Congo—a conflict-affected area marked by extreme poverty. It contributes to the promotion of sexual and reproductive health rights, without discrimination, including the right to information.

Accordingly, this study aims to investigate the types of knowledge that influence both stigma and social support for women seeking safe abortion care. We hypothesize that specific knowledge components contribute significantly to reducing community-level stigma while enhancing social support for women in need of abortion services.

## 2. Methods

### Data Collection and Sample

Data were collected from a sample of 1,715 women and girls of reproductive age, as well as men and boys, as part of a cross-sectional survey conducted from May to June 2024 in the provinces of Kinshasa and North Kivu, regions of the Democratic Republic of Congo (DRC) affected by persistent armed conflict.

Participants were eligible for inclusion if they were at least 16 years old at the time of the survey, mentally capable and willing to provide informed consent, able to speak French, Swahili, or Lingala, and permanently residing in one of the intervention or comparison health zones (i.e., not residing temporarily, seasonally, for work, vacation, or any other short-term reason). Individuals who were not mentally capable of providing informed consent were excluded; interviewers used their judgment to assess whether respondents could understand the consent form and respond appropriately to survey questions.

Among the respondents, only 574 individuals reported knowing at least one woman who had either undergone an abortion or considered terminating a pregnancy. This subsample formed the analytic base for the present study.

### Positionality Statement

This study was conducted by a team of seven researchers, all based in Kinshasa, Democratic Republic of Congo, and of Congolese origin. The group comprises six men and one woman, with professional experience in sexual and reproductive health ranging from four to over twenty years. This diversity in gender and expertise enriched the research process.

The lead author was primarily responsible for the study’s conceptualization, methodological design, data curation, formal analysis, software development, data visualization, and drafting the initial manuscript. The co-authors contributed to supervision, validation, and critical review and editing of the manuscript.

We acknowledge that our social identities, professional backgrounds, and shared geographical context may shape our perspectives and interpretations. To mitigate potential biases, we engaged in continuous reflexivity throughout the research process, documenting and consciously setting aside our assumptions to ensure a rigorous and balanced analysis.

### Measures

Three validated instruments were used for the analysis. Social support was assessed using the Received Social Support Scale (R3S), composed of nine items (SSB1–SSB9). Stigma was measured with the Stigmatizing Attitudes, Beliefs, and Actions Scale (SABAS), comprising 18 items (Q301–Q318). Finally, community knowledge was assessed through four selected items (AK1–AK4) from the Knowledge, Attitudes, Practices, and Intentions (KAPI) survey, originally containing 17 items. All three instruments employed five-point Likert scales (1 = Strongly disagree; 2 = Disagree; 3 = Uncertain; 4 = Agree; 5 = Strongly agree).

### Data Analysis

Given the complexity of interactions between the independent variable (knowledge) and the dependent variables (social support and stigma), and the need to account for latent constructs underlying the psychosocial processes under study, structural equation modeling (SEM) was employed. Analyses were conducted using JASP software.

In the initial model specification, all items were included. To optimize model quality—particularly global fit and construct validity—some items were progressively removed. Item retention was guided by statistical criteria, including overall model fit and reliability. Model adequacy was assessed using the Chi-square test and standard indices: Comparative Fit Index (CFI ≥ 0.95), Tucker-Lewis Index (TLI ≥ 0.95), Root Mean Square Error of Approximation (RMSEA ≤ 0.06), and Standardized Root Mean Square Residual (SRMR ≤ 0.08). Modification indices were considered significant at the threshold of 7.84 (22).

The SABAS, KAPI, and R3S scales have been validated in previous studies (23,24), allowing us to dispense with confirmatory factor analysis (CFA).

### IRB approval Study

The study protocol was approved by the Ethics Committee of the Kinshasa School of Public Health (ESP/CE/087/2024) on April 22^nd^ 2024, and conducted in accordance with international ethical guidelines, including the Declaration of Helsinki and the Belmont Report.

## 3. Results

Table 1 presents socio-demographic characteristics of 574 respondents. The mean age was 26 years (SD = 8), with over half aged 25 or older. Males represented 56.62% of the sample. Education levels were relatively high: 64.82% completed secondary or higher education, though 3.99% had only primary or none.

**Table 1.** Socio-demographic characteristics of the study population (N=574)

| Characteristics | Respondents N=574 |
| --- | --- |
| **Age** |  |
| Mean, SD | 26; SD = 8 |
| **Age group** |  |
| Under 20 years | 19.51% |
| 20 to 24 years | 26.66% |
| 25 years or older | 53.83% |
| **Gender** |  |
| Male | 56.62% |
| Female | 43.38% |
| **Highest level of education** |  |
| None | 0.52% |
| Incomplete primary | 1.39% |
| Completed primary | 2.09% |
| Incomplete secondary | 31.71% |
| Completed secondary | 37.11% |
| Higher/University | 27.18% |
| **Main occupation** |  |
| No occupation/Housewife/Student or pupil | 47.91% |
| Public sector employee with regular monthly salary | 8.01% |
| Private sector employee with regular monthly salary | 5.05% |
| Self-employed in the private sector (liberal) | 17.42% |
| Informal sector worker and small trade | 20.56% |
| Agro-pastoral and fishing | 0.17% |
| Other | 0.87% |
| **Marital status** |  |
| Married | 21.95% |
| Living with partner, not married | 7.84% |
| In a stable relationship, not living together | 4.18% |
| No regular partner / single | 63.76% |
| Separated/divorced | 1.57% |
| Widowed | 0.52% |
| Refused to answer | 0.17% |

Nearly half (47.91%) reported no formal occupation, while informal work (20.56%) and self-employment (17.42%) were common; only 13.06% held regular salaried jobs. Most participants were single (63.76%), while 21.95% were married.

Table 2 presents the baseline reliability indices of the estimated model. The null hypothesis, that the empirical covariance matrix is equivalent to the hypothesized model covariance matrix, was not rejected (χ² = 33.028, df = 31, p = 0.368). This non-significant result indicates a satisfactory model fit.

**Table 2.**
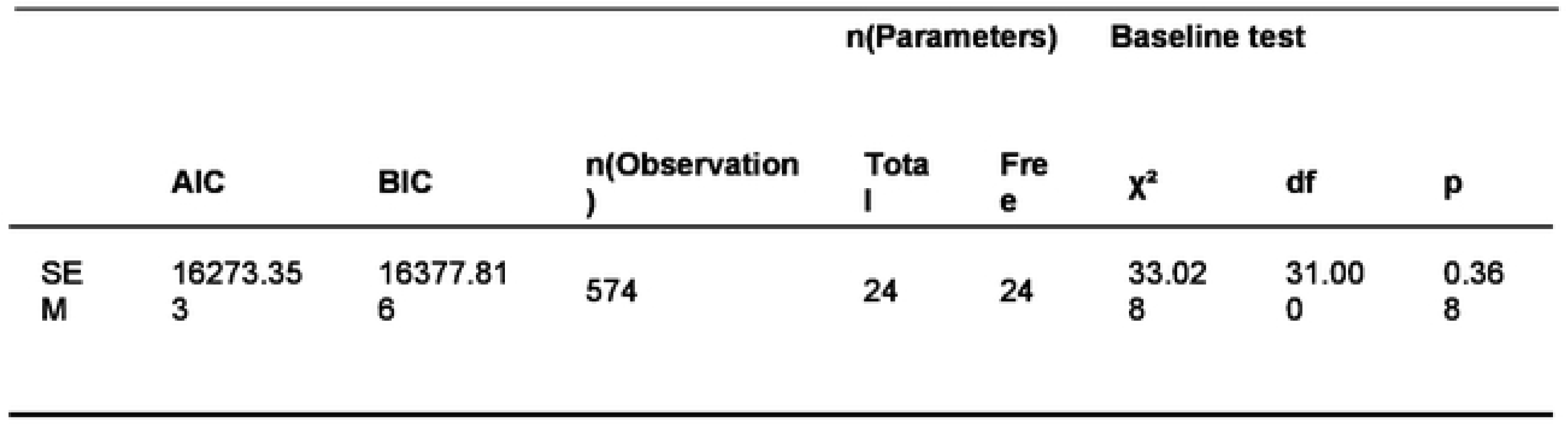
Model fit indices.

Incremental fit indices were excellent, with both the Comparative Fit Index (CFI = 0.999) and the Tucker-Lewis Index (TLI = 0.999) exceeding the recommended threshold of 0.95. Similarly, absolute fit indices confirmed robust model adequacy, with RMSEA = 0.011 and SRMR = 0.025, both well below the accepted cut-offs of 0.06 and 0.08, respectively.

As shown in Table 3, the knowledge construct accounted for 9.3% of the variance in social support (SocialSup) and 3.6% of the variance in stigma (Stigm) within the community. While modest, these proportions indicate a non-trivial contribution of knowledge to psychosocial outcomes. At the item level, several indicators (e.g., SSB3, SSB4, q315, q316) exhibited strong explanatory power, with R² values ranging between 0.71 and 0.89.

**Table 3.** R-squared values.

|  | R<sup>2</sup> |
| --- | --- |
| SSB3 | 0.779 |
| SSB4 | 0.888 |
| SSB5 | 0.608 |
| SSB7 | 0.356 |
| AK1 | 0.277 |
| AK2 | 0.654 |
| q301 | 0.224 |
| q302 | 0.390 |
| q315 | 0.712 |
| q316 | 0.745 |
| SocialSup | 0.093 |
| Stigm | 0.036 |

In Table 4, half of the indicators demonstrated loadings above the 0.70 threshold, reflecting strong construct representation. Among stigma-related items, q315 (β = 0.844, 95% CI: 0.801–0.886, p<0,001) and q316 (β = 0.863, 95% CI: 0.822–0.905, p<0,001) showed particularly robust associations. Similarly, the social support items SSB3 (β = 0.883, 95% CI: 0,858–0.907, p<0,001), SSB4 (β = 0.943, 95% CI: 0,922–0.963, p<0,001), and SSB5 (β = 0.780, 95% CI: 0,744–0.816, p<0,001) were among the most reliable indicators.

**Table 4.**
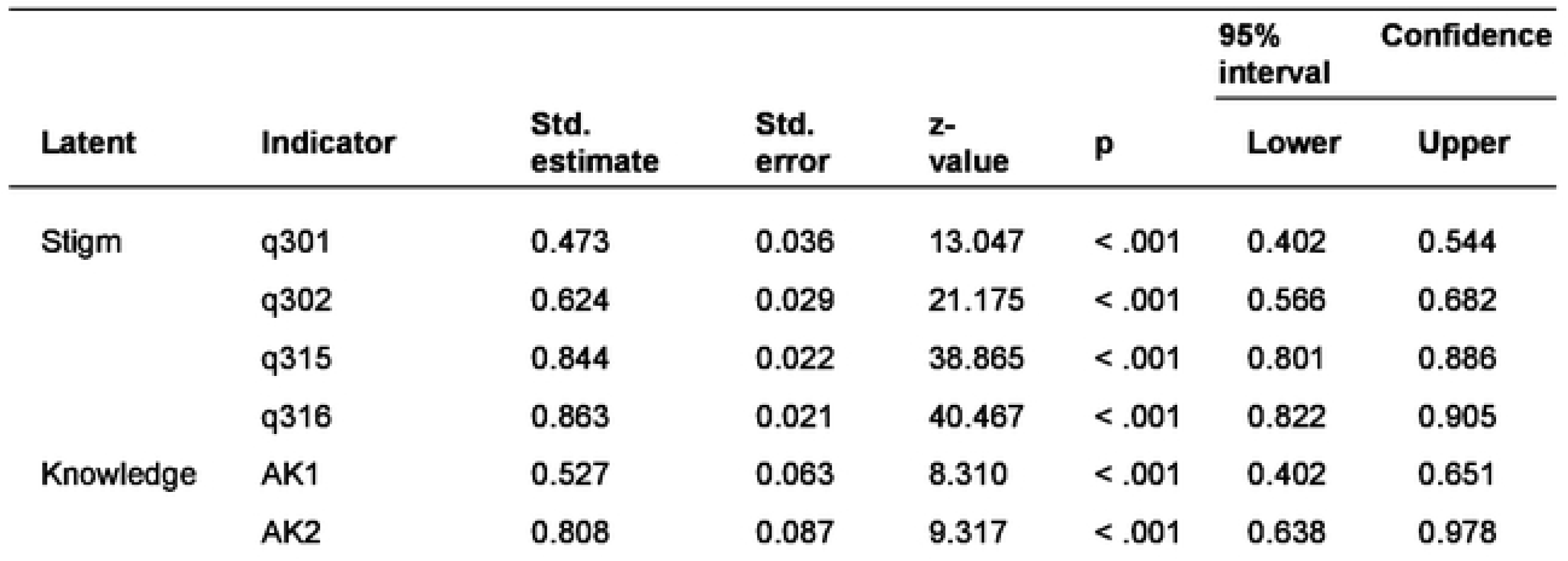
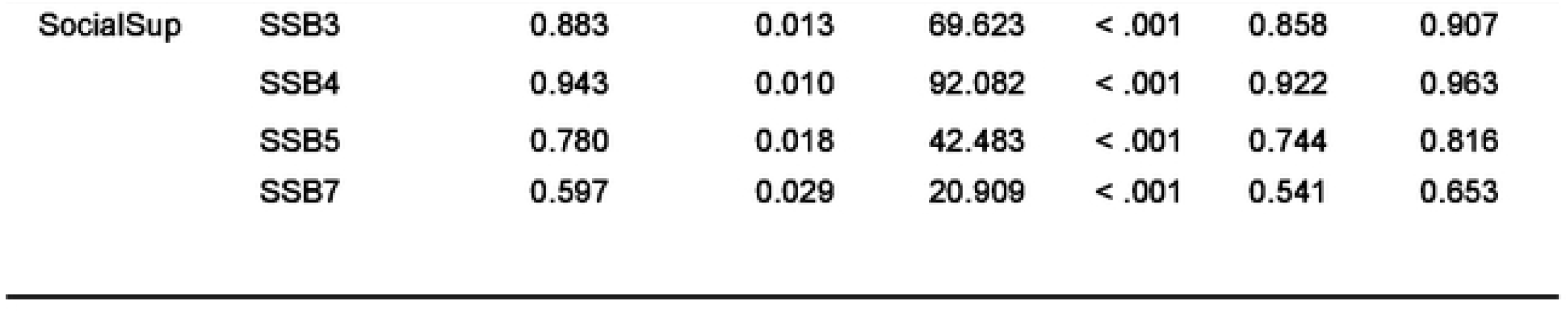
Factor loadings.

Residual variances, are presented in Appendix B (see Table B1), fell within acceptable ranges, suggesting that the retained items contributed meaningfully to their respective latent constructs. For example, residuals for SSB3 (Std. Est.=0.221, 95% CI: 0,117–0.265, p< 0,001) and SSB4 (Std. Est.= 0.112, 95% CI: 0,074–0.149, p<0,001) were relatively low, indicating high item reliability. By contrast, items AK1 (Std. Est.= 0.723, 95% CI: 0,592–0.853, p<0,001) and q301 (Std. Est.= 0.776, 95% CI: 0,709–0.844, p<0,001) retained higher residual variance, signaling areas of weaker measurement precision.

Additionally, covariance analyses revealed an association between stigma items q301 and q302. Accounting for this covariance improved the overall model fit, further supporting the robustness of the final specification.

As summarized in Table 5, knowledge exerted a positive total effect on social support and a negative total effect on stigma.

**Table 5.** Total effects.

|  | Std.<br>estimate | Std.<br>Error | z-value | p | 95%<br>interval | Confidence |
| --- | --- | --- | --- | --- | --- | --- |
|  |  |  |  |  | Lower | Upper |
| Knowledge → Stigm | -0.191 | 0.062 | -3.084 | 0.002 | -0.312 | -0.070 |
| Knowledge→SocialSup | 0.304 | 0.056 | 5.444 | < .001 | 0.194 | 0.413 |

Consistent with these results (see Table 5), improved community knowledge was associated with enhanced social support for women seeking abortion care (Std. Est.= 0,304, 95% CI: 0,194 – 0,413, p < 0.001). As shown in Table 5, higher levels of knowledge were linked to a measurable reduction in stigmatizing attitudes, beliefs, and behaviors toward women who undergo abortion (Std. Est.= - 0,191, 95% CI: -0,312 –-0,070, p = 0.002). The standardized estimates presented in Table 5 indicate that knowledge exerts a moderate effect on social support (Std. Est. = 0.304 > 0.30), whereas its effect on stigma, although statistically significant, remains relatively weak (Std. Est.= −0.191 < 0.30).

As summarized in Figure 1, The path diagram illustrates the central role of knowledge in shaping both social support and stigma within the community. Specifically, two knowledge indicators—AK1 (awareness of the legal conditions under which abortion is permitted) and AK2 (understanding that abortion is a safe medical procedure)—emerge as pivotal drivers of the psychosocial dynamics observed.

**Figure 1.**
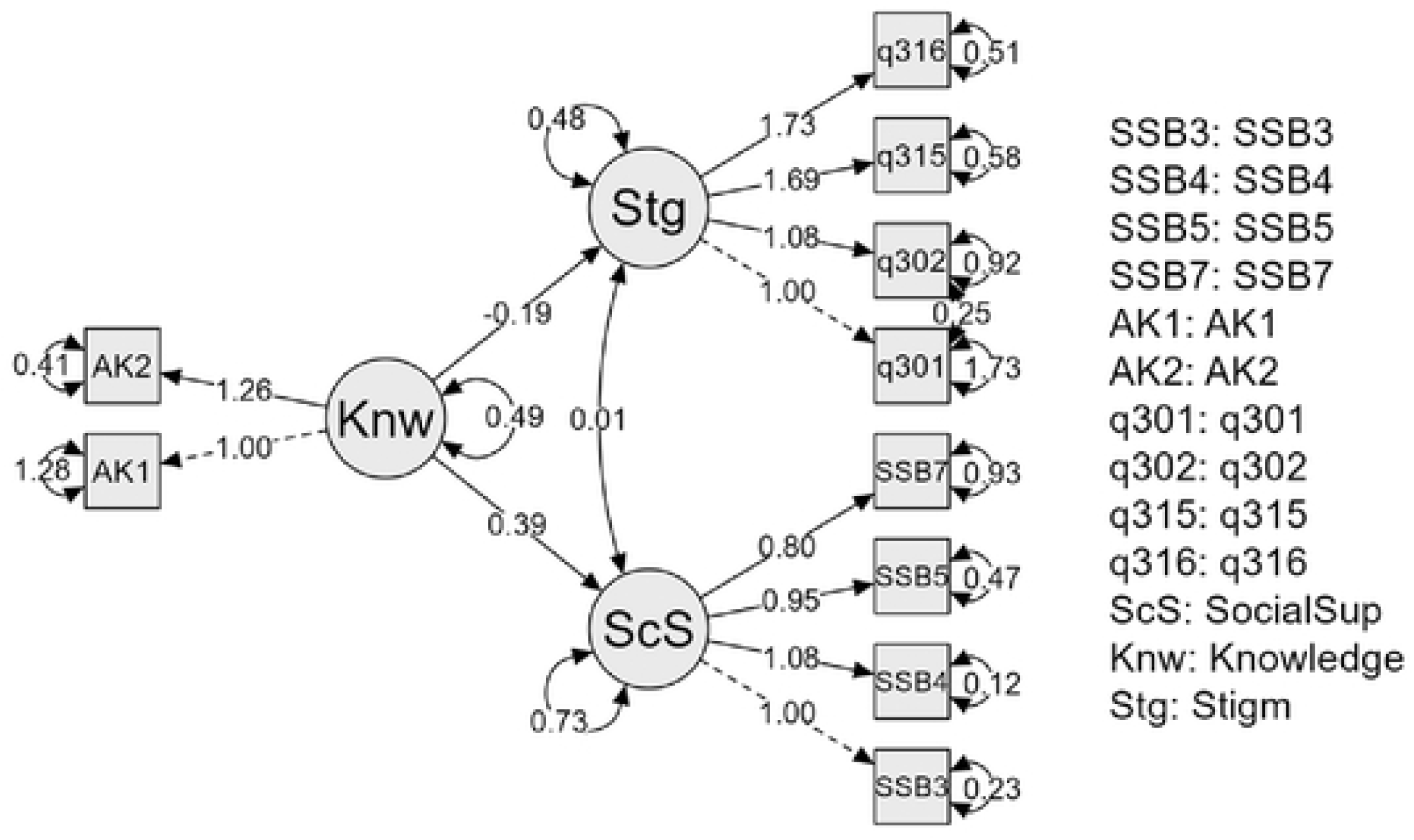
Path diagram of the structural equation model

On the one hand, these knowledge items contribute to reducing stigmatizing attitudes and beliefs. Greater awareness was associated with a rejection of notions such as the belief that women who undergo abortion should be excluded from religious services (q301), that abortion is inherently sinful (q302), that women who terminate pregnancies are “bad mothers” (q315), or that they bring shame to their community (q316).

On the other hand, knowledge significantly enhances manifestations of social support. It fosters empathy toward women seeking abortion (SSB3), encourages willingness to provide financial assistance for abortion-related care (SSB4), promotes openness to listening to women’s concerns about pregnancy (SSB5), and strengthens support in decision-making regarding pregnancy outcomes (SSB7).

Overall, a one-unit increase in the knowledge score was associated with a 0.39-point increase in social support and a 0.19-point reduction in stigma. These findings underscore that community knowledge serves as a dual lever, simultaneously reinforcing supportive behaviors while mitigating stigmatizing attitudes toward women seeking abortion care.

## 4. Discussions

The findings of this study corroborate the initial hypothesis that improved knowledge, specifically regarding the legal conditions for accessing abortion services and the safety of the procedure when performed by qualified health professionals, serves as a crucial lever for reducing stigma and strengthening social support for women seeking abortion care. These results confirm that social dynamics surrounding abortion are inherently interdependent.

Our analyses revealed a direct positive effect of knowledge on social support (Std. Est.= 0,304, 95% CI: 0,194 – 0,413, p < 0.001), aligning with prior evidence from other settings. Bekalu et al. (2022) demonstrated that accurate information and community awareness of sexual and reproductive health foster empathy and solidarity toward women facing unintended pregnancies (20). Similarly, Seymour et al. (2023) underscored that access to informational and decisional support is a major driver of enhanced social and emotional support throughout the abortion trajectory (10). Furthermore, Cockrill et al. (2013) and Hanschmidt et al. (2016) highlighted that greater knowledge of abortion contributes to reducing negative attitudes, thereby creating a more favorable climate for social support (25,26). Nonetheless, as Kassie et al. (2020) noted, knowledge alone is not always sufficient to transform entrenched social norms unless accompanied by structural and policy-level interventions (27).

With regard to stigma, our findings show that increased knowledge contributes to reducing stigmatizing attitudes (Std. Est.= - 0,191, 95% CI: -0,312 – -0,070, p = 0.002), although this effect remains modest. This observation is consistent with LeTourneau (2016), who argued that the persistence of rigid religious or moral beliefs can limit the impact of knowledge alone, particularly in contexts where stigma is deeply entrenched (21). Qualitative studies conducted in the DRC and other low- and middle-income countries (Burtscher et al., 2020; Casey et al., 2019) similarly confirmed that women who undergo abortion are frequently perceived as sources of shame and subjected to social exclusion, despite the existence of favorable policy frameworks such as the Maputo Protocol(15,16).

Our findings also align with broader literature emphasizing the protective role of social support. Evidence from the United States and Latin America (Rocca et al., 2015; Bercu et al., 2022) has shown that the quality of interpersonal and social support not only shapes women’s emotional experiences but also influences their ability to access safe health services (11,28). In the African context, studies in Uganda and Malawi (Lubinga et al., 2013; Levandowski et al., 2012) highlighted that the absence of social support exacerbates medical complications and psychological consequences associated with unsafe abortions (29,30).

### Strengths and Limitations

A major strength of this study lies in its analytical framework, which simultaneously integrates knowledge, stigma, and social support, thereby enabling the examination of their complex interactions in the context of the Democratic Republic of Congo. Furthermore, the use of validated measurement tools—specifically the Received Social Support Scale (R3S) and the Stigmatizing Attitudes, Beliefs, and Actions Scale (SABAS)—enhances the robustness and credibility of the findings.

Nonetheless, several limitations warrant consideration. First, the study did not incorporate demographic variables or key contextual factors that may influence levels of stigma and social support. Second, given the sensitivity of the topic, the possibility of social desirability bias in participants’ responses cannot be excluded. Finally, as the findings are embedded within the socio-cultural and political realities of the DRC, caution must be exercised when generalizing them to other contexts.

## 5. Conclusion

This study highlights the decisive role of knowledge in reshaping the psychosocial dynamics surrounding abortion within communities. Specifically, two key dimensions of knowledge, awareness that abortion can be a safe medical procedure and understanding the legal conditions under which it is permitted, emerged as powerful levers for reducing stigma and strengthening social support.

Our findings underscore the need to intensify community-based communication and educational strategies aimed at improving awareness of abortion and its public health implications. At the same time, targeted stigma-reduction interventions are essential, particularly those engaging community leaders, religious authorities, and healthcare providers as agents of change.

From a policy perspective, the results suggest that combining structural reforms with educational initiatives could significantly enhance social support, thereby facilitating greater access to comprehensive and safe abortion care in the Democratic Republic of Congo.

## Data Availability

All data used are integreted

## Author Contributions

Conceptualization : Frédéric Etsou

Data curation : Frédéric Etsou, Bergitho Bokamba

Formal analysis: Frédéric Etsou

Methodology : Frédéric Etsou

Software : Frédéric Etsou, Bergitho Bokamba

Supervision : Nadia Lobo

Validation : Jean-Claude Mulunda, Mike Mpoyi

Visualization: Frédéric Etsou

Writing - original draft Writing: Frédéric Etsou

Writing - Review & editing: Michée Kanda, Joseph Ngwanza, Mike Mpoyi, Jean-Claude Mulunda, Nadia Lobo

## Acknowledgements

We sincerely thank principal investigator, **Professor Eric Mafuta**, Director of the Kinshasa School of Public Health, for supervising the management and transmission of all survey data. We also extend our gratitude to the entire team of data collectors and the Youthsprint Coalition for their active involvement in the data collection process. Their dedication and support were essential to the successful completion of this study.

## Ethics Declarations

Electronically informed consent was obtained from all participants, who were informed about study objectives, procedures, potential risks and benefits, and their right to withdraw at any time. Confidentiality and anonymity were maintained throughout.

This research involved adult participants and complied with all legal and ethical requirements in the Democratic Republic of Congo. No participants are identifiable in this manuscript.

ChatGPT (GPT-5) was used solely to translate content from French to English. All translations were reviewed and verified by the authors. AI was not involved in data analysis, interpretation, or drafting of the manuscript

## Abbreviations

DHS: Demographic Health Survey
CAC: Comprehensive Abortion Care
DRC: Democratic Republic of Congo
SDMPR: Surveillance de Décès Maternels, périnatals et Riposte
WHO: World Health Organization
R3S: Received Social Support Scale
SABAS: Stigmatizing Attitudes, Beliefs and Actions Scale
SEM: Structural Equations Modelling
CFA: Confirmatory factor analysis
CFI: Comparative fit index
TLI: Tucker-Lewis Index
RMSEA: Root mean square error of approximation
SRMR: Standardized root mean squared residuals
KAPI: Knowledge, Attitudes, Practices, and Intentions

## APPENDIXES

APPENDIX A. Syntax within JASP software version 0.19.3.0

\# measurement model

SocialSup =∼ SSB3 + SSB4 + SSB5 + SSB7

Knowledge=∼ AK1+ AK2

Stigm =∼ q301 + q302 + q315 + q316

#Regression

Stigm ∼ Knowledge

SocialSup ∼ Knowledge

#Index

q301∼∼q302

## Footnotes

1 Les résultats de ce diagramme ne sont pas standardisés

